# Humoral and cellular immune responses and their kinetics vary in dependence of diagnosis and treatment in immunocompromised patients upon COVID-19 mRNA vaccination

**DOI:** 10.1101/2021.12.13.21267603

**Authors:** A. Wagner, E. Garner-Spitzer, A. Schötta, M. Orola, A. Wessely, I. Zwazl, A. Ohradanova-Repic, G. Tajti, L. Gebetsberger, B. Kratzer, E. Tomosel, M. Kutschera, S. Tobudic, W. F. Pickl, M. Kundi, H. Stockinger, G. Novacek, W. Reinisch, C Zielinski, U. Wiedermann

**Author notes:** **Corresponding Author:** Prof. Ursula Wiedermann, MD, PhD, Institute of Specific Prophylaxis and Tropical Medicine, Centre for Pathophysiology, Infectiology and Immunology, Medical University of Vienna, Kinderspitalgasse 15, Vienna A-1090, Austria.

## Abstract

**Background:** Knowledge about humoral and cellular immunogenicity and their kinetics following SARS-CoV-2 mRNA vaccinations in immunosuppressed patients is limited.

**Methods:** Antibody and cytokine responses were assessed in 263 patients with either solid tumors (SOT, n=63), multiple myeloma (MM, n=70) or inflammatory bowel diseases (IBD, n=130) undergoing various immunosuppressive regimens and from 66 healthy controls before the first and the second, as well as four weeks and 5-6 months after the second mRNA vaccine dose with either BNT162b2 or mRNA-1273.

**Findings:** Four weeks after the second dose, seroconversion was lower in cancer than in IBD patients and controls, with the highest non-responder rate in MM patients (17.1%). S1-specific IgG levels correlated with neutralizing antibody titers. While antibody responses correlated with cellular responses in controls and IBD patients, IFN-γ and antibody responses did not in SOT and MM patients. At six months, 19.6% of patients with MM and 7.3% with SOT had become seronegative, while IBD patients and controls remained seropositive in 96.3% and 100%, respectively. Vaccinees receiving mRNA-1273 presented higher antibody levels than those vaccinated with BNT162b2.

**Interpretation:** Cancer patients may launch an inadequate seroresponse in the immediate time range following vaccination and up to six months, correlating with vaccine-specific cellular responses. These findings propose antibody testing in immunosuppressed - along with cellular testing - provides guidance for administration of additional vaccine doses, or may indicate the necessity for antibody treatment. IBD patients respond well to the vaccine, but treatment such as with TNF-α inhibitors may reduce persistence of immune responses.

**Funding:** The study was sponsored and financed by the Medical University of Vienna – third party funding by the Institute of Specific Prophylaxis and Tropical Medicine. AOR. and HS acknowledge funding by the Austrian Science Fund (FWF, P 34253-B).

## Introduction

Infections with the SARS-CoV-2 virus may lead to severe disease in the immunosuppressed population.^1,2^ COVID-19 vaccines have been developed to combat the ongoing pandemic by preventing disease and particularly severe courses of disease. These vaccines have been shown to be highly immunogenic in healthy individuals.^3,4^ However, immunogenicity and vaccine efficacy in patients under different immunosuppressive/modulatory treatments is still a matter of concern. Humoral and/or cellular immune responses are generally affected to different degrees depending on the underlying disease and treatment regimens, which determine the grade of immunosuppression. Thus, impaired vaccine responses have been described for cancer patients in particular under chemotherapy and with progressive disease as well as with low lymphocyte counts around four weeks after the second vaccine dose.^5-7^ With regard to inflammatory bowel disease (IBD), it has been shown that the type of treatment - particularly with TNF-α blockers - results in decreased immunogenicity, when measured up to 2 weeks after the second vaccine dose, as compared to healthy individuals, although shown in an only small cohort.^8^

Magnitude of response may also be related to the type of mRNA vaccine used, as for patients immunosuppressed by anti-CD20 therapy or with multiple myeloma better seroconversion rates have been reported following the administration of mRNA-1273 (Moderna) rather than of BNT162b2 (Pfizer BioNTech) vaccines.^6,9^ Very little data is available regarding the vaccine-specific cellular responses and whether a correlation or dissociation of humoral and cellular immune response can predict responsiveness or non-responsiveness to the vaccine and subsequent protection or failure to protect against disease. Another unanswered question is the persistence of immune responses in seropositive immunosuppressed patients upon two doses of mRNA vaccines.

Here, we present data obtained from patients receiving different immunosuppressive/-modulatory treatment regimens including cancers (solid tumors (SOT) and multiple myeloma (MM)) as well as IBD compared to healthy control individuals without previous COVID-19 infection. We characterized humoral and cellular responses after two doses of COVID-19 mRNA vaccines in relation to their underlying leukocyte distribution in order to find potential predictors for vaccine (non-)responsiveness to this neo-antigen. We also aimed to investigate whether humoral (non-)responsiveness is linked to cellular responsiveness to provide a better estimation of the level of vaccine responsiveness and remaining infection susceptibility. Furthermore, we differentiated between BNT162b2- and mRNA-1273-vaccinated individuals in order to estimate if a favourable use of one over the other vaccines might be considered in relation to the severity of immunosuppression. Finally, we evaluated the persistence of antibody levels over a period of six months before participants were eligible for a third vaccine dose and related these findings with the currently existing recommendations.

## Methods

### Study population

We included in total 329 participants with underlying SOT, MM or IBD and under various immunosuppressive/ -modulatory treatment as well as healthy controls in the finals analysis (Suppl Fig 1 and 2 and Suppl materials). We obtained written informed consent from all patients according to the Declaration of Helsinki/International Conference on Harmonisation Guideline for Good Clinical Practice. The study was approved by the Ethics Committee of the Medical University of Vienna (EK: 1073/2021).

### Humoral responses

SARS-CoV-2-specific IgG antibodies directed against the subunit 1 (S1) of the spike protein were measured by ELISA (Quantivac®, Euroimmun) in diluted serum samples (1:101) according to the manufacturer’s instructions in all participants. Antibody quantification results are expressed in binding antibody units/ml (BAU/ml) and values above 35.2 BAU/ml were considered as positive as defined by the manufacturer.

Neutralization assay was performed in the preselected subgroup of patients with the human SARS-CoV-2 isolate BetaCoV/Munich/BavPat1/2020 kindly provided by Christian Drosten, Charité, Berlin, and distributed by the European Virology Archive (Ref-SKU: 026V-03883).^10^ The assay was performed according to the protocol by Amanat et al. (for detailed information please see Suppl materials).^11^

### Leukocyte phenotyping

Phenotyping of leukocyte subpopulations in whole blood samples was performed by flow cytometric analysis after staining with directly-conjugated monoclonal antibodies, as previously described^12^ and absolute cell counts were calculated.

### T-cell responses

PBMCs were isolated from heparinized human peripheral blood via density gradient centrifugation using Ficoll™ (LSM 1077, PAA, Pasching, Austria) as previously described.^13^ Isolated PBMCs were rested over night before stimulation with peptide pool covering either the S1 domain of the SARS-CoV-2-specific spike protein or the nucleocapsid protein for 24 hours. Supernatants were stored at -80°C until analysis using Luminex® 100/200 System to determine concentration of interleukin (IL)-2, IL-5, IL-10, IL-17a, IL-22, granulocyte-macrophage colony-stimulating factor (GM-CSF) and interferon (IFN)-γ (for detailed information see Suppl Materials).

### Statistical analysis

Seroprevalence data are presented as descriptive statistics. To evaluate differences between the different study groups, we compared results with non-parametric Kruskal-Wallis test including Dunn’s multiple comparisons test. Correlations were calculated by linear regression analysis. P-values <0.05 were considered significant. Comparison of cytokine production before the first and after the second dose and decrease of antibody levels between four weeks and five to six months after the second vaccine dose were calculated by paired t-test.

## Results

### Demographic data

Between March 2021 und June 2021, 263 patients (55.9 % females, mean age 54.0 ± 15.2) and 66 controls (50% females, mean age 46.1 ± 15.1) were included in the study. Patients had either SOT (n=63; 83.3% females, mean age 62.8 ± 11.5) of the breast (71%) or lung (29%), MM (n=70; 44.3% females, mean age 66.5 ± 8.0) or IBD (n=130; 50.0% females, mean age 46.1 ± 15.1) and were undergoing different treatment regimens (Table 1 and 2).

**Table 1:** Description of the study population (n=263 patients, n=66 controls)

|  | SOT (n=63) | MM (n=70) | IBD (n=130) | controls (n=66) |
| --- | --- | --- | --- | --- |
| Age (y) mean ± SD; (range) | 62.8 ± 11.5;<br>(38-82) | 66.5 ± 8.0;<br>(46-83) | 44.0 ± 14.4;<br>(19-77) | 46.1 ± 15.1;<br>(20-78) |
| mean BMI ± SD | 24.8 ± 5.0 | 26.1 ± 4.0 | 31.4 ± 6.7 | 25.5 ± 4.5 |
| n= of obese<br>(BMI > 30) | 7 | 12 | 12 | 12 |
| Gender |  |  |  |  |
| Female n (%) | 55 (87.3) | 31 (44.3) | 61 (46.9) | 33 (50.0) |
| Mean time from diagnosis (y) | 5.3 ± 7.1 | 8.3 ± 6.9 | 15.0 ± 10.7 | n.a. |
| Vaccine |  |  |  |  |
| BNT162b2 n (%) | 36 (57.1) | 48 (68.6) | 2 (1.5) | 5 (7.6) |
| mRNA-1273 n (%) | 27 (42.9) | 22 (31.4) | 128 (98.5) | 61 (92.4) |
| Ongoing immunosuppressive/-<br>immunomodulator treatment n (%) | 49 (77.8) | 45 (64.3) | 111 (83.8) | n.a. |
| Stem cell transplantation n (%) | n.a. | 32 (45.7) | n.a. | n.a. |

n.a. (not applicable)

**Table 2.** Characteristics of IBD patients.

| IBD type | n (%) | Gender<br>f: n (%) | Age mean ± SD<br>(range) | Time from<br>diagnosis in years ±<br>SD | Ongoing immuno-<br>suppressive/ -modulatory<br>treatment n (%) |
| --- | --- | --- | --- | --- | --- |
| Total | 130 | 61 (46.9) | 19-77 | 15.0 ± 10.7 | 111 (83.8) |
| Crohn's disease (CD) | 81 (62.3) | 36 (44.4) | 44.0 ± 12.8 (19-77) | 16.4 ± 10.5 | 70 (86.4) |
| Ulcerative colitis (UC) | 48 (36.9) | 25 (52.1) | 44.6 ± 16.6 (20-74) | 13.3 ± 10.7 | 39 (81.3) |
| IBD unclassified (IBDU) | 1 (0.8) | 0 (0) | 23.0 ± 0.0 (23) | 3.0 ± 0.0 | 0 (0%) |

### Humoral immune responses

S1-specific IgG antibody levels were determined by ELISA before the first and the second dose and 4 weeks as well as 5-6 months after the second dose.

After the first dose, 50.0% of MM patients, 28.6% of SOT patients and 3.8% of IBD patients had a negative antibody result, as compared to only 1.5% of the control (Fig 1a). After the second dose, the non-responder rate dropped to 17.1% in MM patients and 1.6% in SOT patients with different treatment regimens (Table 3), whereas all patients with IBD and controls showed a positive antibody result.

**Figure 1.**
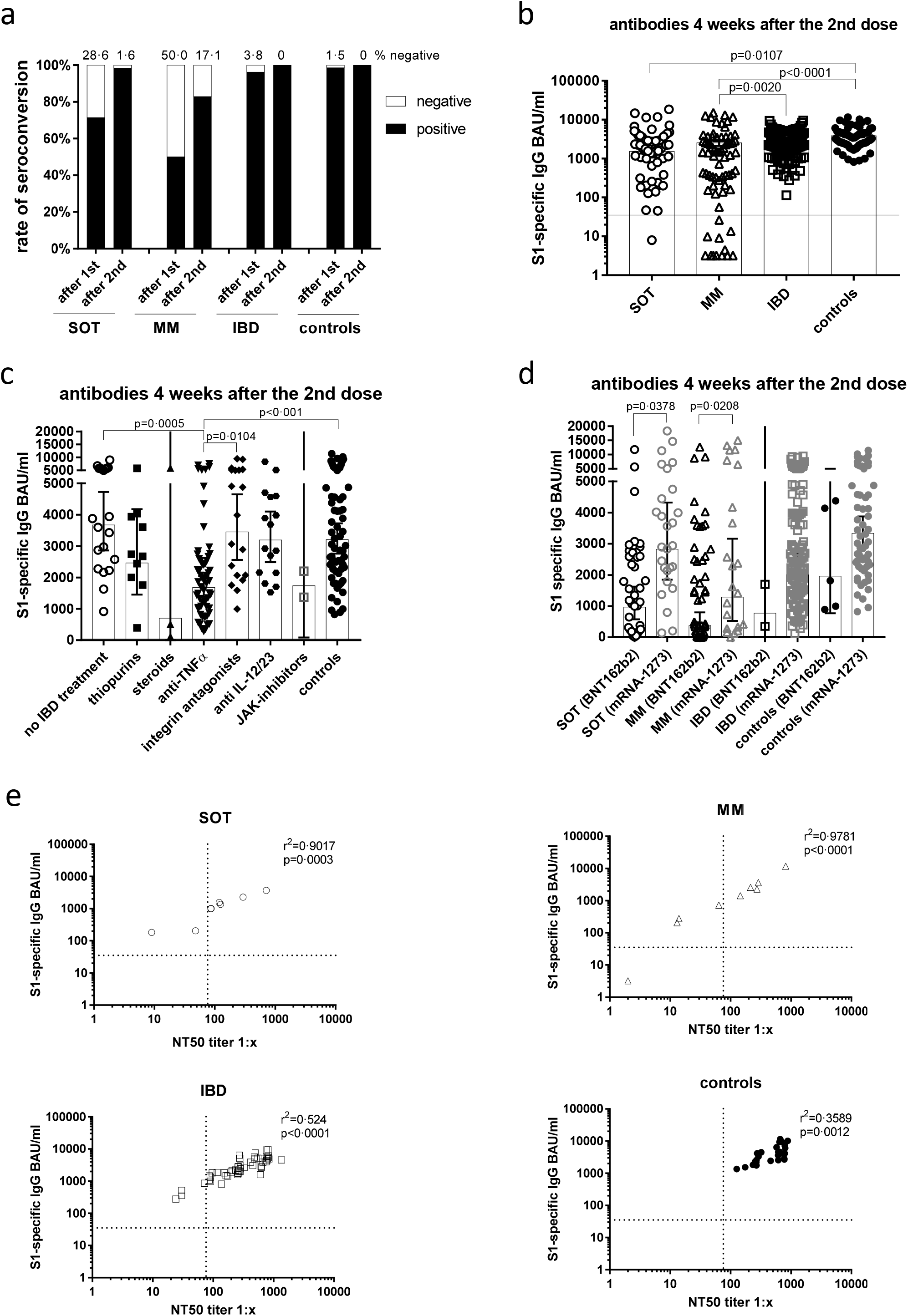
Antibody responses four weeks after the second mRNA vaccination and correlation of S1-specific IgG with neutralizing antibodies. (a) Seroconversion rates after the first and second dose in all study participants of all groups. (b) Individual S1-specific IgG levels of all participants. Differences between the groups below p values of 0.05 were regarded as significant. The black line represents the threshold for positive results (35.2 BAU/ml). (c) S1-specific IgG levels of IBD patients in respect of their treatment and in comparison to the controls. (d) S1-specific antibody levels in relation to the type of mRNA vaccine applied (BNT162b2 or mRNA-1273), whereby due to the number of participants statistical differences could only be calculated for SOT and MM patients. (e) Correlation of S1-specific IgG levels with the NT50 of a neutralization test with sera taken four weeks after the second vaccination. Differences between the groups below p values of 0.05 were regarded as significant. Dotted lines indicate the threshold for positive results (35.2 BAU/ml). Columns represent GMC with 95% confidence interval (CI).

**Table 3.**
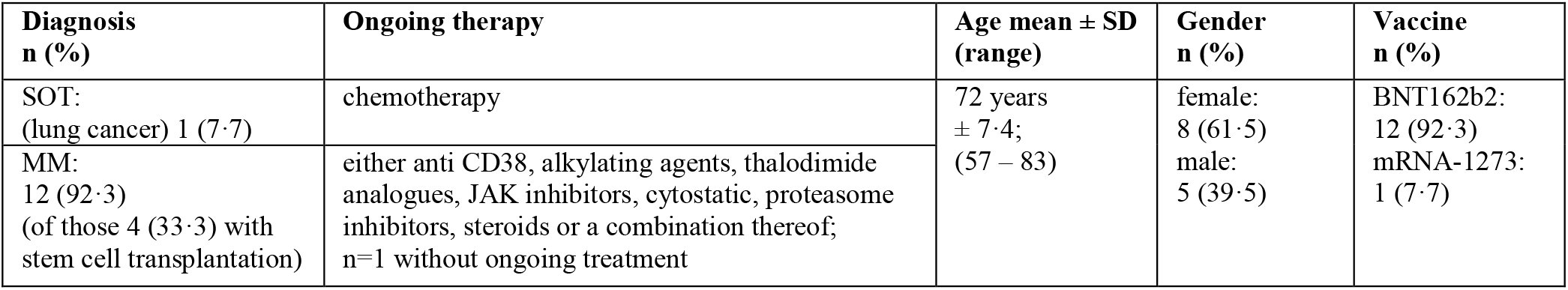
Characteristics of non-responders at four weeks after the second dose. Clinical and demographic parameters of those 13 patients with negative and borderline antibody responses measured at four weeks after the second vaccine dose.

| Diagnosis<br>n (%) | Ongoing therapy | Age mean ± SD<br>(range) | Gender<br>n (%) | Vaccine<br>n (%) |
| --- | --- | --- | --- | --- |
| SOT:<br>(lung cancer) 1 (7.7) | chemotherapy | 72 years<br>± 7.4;<br>(57 – 83) | female:<br>8 (61.5)<br>male:<br>5 (39.5) | BNT162b2:<br>12 (92.3)<br>mRNA-1273:<br>1 (7.7) |
| MM:<br>12 (92.3)<br>(of those 4 (33.3) with<br>stem cell transplantation) | either anti CD38, alkylating agents, thalodimide<br>analogues, JAK inhibitors, cytostatic, proteasome<br>inhibitors, steroids or a combination thereof;<br>n=1 without ongoing treatment |  |  |  |

**Table 4.** Characteristics of participants that became seronegative at five to six months after the second dose. Clinical and demographic parameters of those 19 patients with negative and borderline antibody responses measured at five to six months after the second vaccine dose.

| Diagnosis<br>n (%) | Ongoing therapy | Age mean ± SD<br>(range) | Gender<br>n (%) | Vaccine<br>n (%) |
| --- | --- | --- | --- | --- |
| SOT:<br>4 (21.0)<br>[lung cancer: 2 (10.5);<br>breast cancer: 2 (10.5)] | antimetabolites, alkylating agents, anti-PD-1,<br>mitotic inhibitors, hormones, steroids | 63 ± 12.9;<br>(31–81) | Female:<br>8 (42.1)<br>male: 1<br>1 (57.9) | BNT162b2:<br>10 (52.6),<br>mRNA-1273:<br>9 (47.4) |
| MM:<br>11 (57.9)<br>(of those 6 with stem cell<br>transplantation) | thalidomide analogue, proteasome inhibitor,<br>steroids, anti-CD38;<br>n=3 without ongoing treatment |  |  |  |
| IBD:<br>4 (21.1) [CD 3 (15.8);<br>UC 1 (5.3) ] | thiopurins, TNF-α blockers, steroids | | | |

When comparing antibody levels measured four weeks after the second dose, the lowest geometric mean concentrations (GMC) were reached in MM patients (552.5 GMC), as compared to IBD (2275.3 GMC; p=0.0020) and controls (3205.5 GMC; p<0.0001) (Fig 1b). Within the group of SOT patients (1529 GMC), antibody levels were significantly lower than in controls (3205.5 GMC; p=0.0107) with lower titers in the lung cancer (694.9 GMC) than in the breast cancer group (2096.4 GMC; p=0.0061). With regard to age, only SOT patients below 60 years displayed higher S1-specific antibody levels (p=0.0052) than those 60 years and older (Suppl Figure 3a). No significant differences in antibody levels between female and male participants were detected within the mentioned groups (Suppl Fig 3b).

Subgroup analysis of IBD patients showed that four weeks after the second dose, patients who were treated with TNF-α inhibitors had significantly lower antibody levels (n= 59; 1685 GMC) than IBD patients without current immunosuppressive/-modulatory medication (n=21; 3676 GMC; p=0.0005), patients receiving the α4β7 integrin antagonist vedolizumab (n=19; 3454 GMC; p=0.0104) or controls (n=66; 3206 GMC; p<0.001) (Fig 1c). The decreased antibody levels were found with all TNF-α inhibitors (infliximab: n=16, 1319 GMC; adalimumab: n= 38, 1799 GMC; golimumab: n=5; 2246 GMC) without significant differences between the three compounds.

Subanalysis of antibody titers according to the mRNA vaccine used revealed that in SOT as well as in MM patients, antibody titers were higher after vaccination with mRNA-1273 (SOT: 2827 GMC, MM: 1289 GMC) than with BNT162b2 (SOT: 964.5 GMC, MM: 374.8 GMC; p<0.05). With regard to non-responders, all (except one) were vaccinated with BNT162b2 (Table 3). For IBD and controls, there were only two and five individuals, respectively, who were vaccinated with BNT162b2 not allowing for appropriate comparisons of outcome (Fig 1d).

We further evaluated neutralization titers in a live-virus assay in 28.9% of the study participants (those in the subgroup for cellular analysis) and found a good correlation of S1-specific IgG measured by ELISA in all groups (p<0.01) (Fig 1e). Furthermore, we analysed the neutralization titer prior to the first and four weeks after the second vaccine dose. None of the participants showed a positive neutralization titer before vaccination. After vaccination the geometric mean titers (GMT) were significantly lower in SOT (n=8; 99.9 GMT; p=0.0044) and MM patients (n=9; 71.8 GMT; p=0.0073) compared to controls (n=25; 453.3 GMT) (Suppl Fig 3c). An additional analysis of ELISA-negatives in the neutralization test demonstrated that neutralizing antibodies were also not detectable (Suppl Fig 3d).

### Cellular immune responses

In a preselected subgroup of participants, we analyzed the T cell response by using a cytokine release assay. After the second vaccine dose, the immune system of the IBD patients and the controls, mounted a clear T cell response upon stimulation with the peptide pool of the S1 subunit of the SARS-CoV-2 spike protein. The T cells secreted the T cell growth factor IL-2, the pro-inflammatory cytokines IFN-γ, IL-17a and GM-CSF, the regulatory cytokine IL-10 as well as the Th2 cytokine IL-5. The T cells of the SOT patients presented with the ability to secrete IL-2, IFN-γ, IL-10, IL-17a and GM-CSF, whereas only IFN-γ and concomitant IL-10 and IL-17a were induced in MM patients (Fig 2 and Suppl Fig 4).

**Figure 2.**
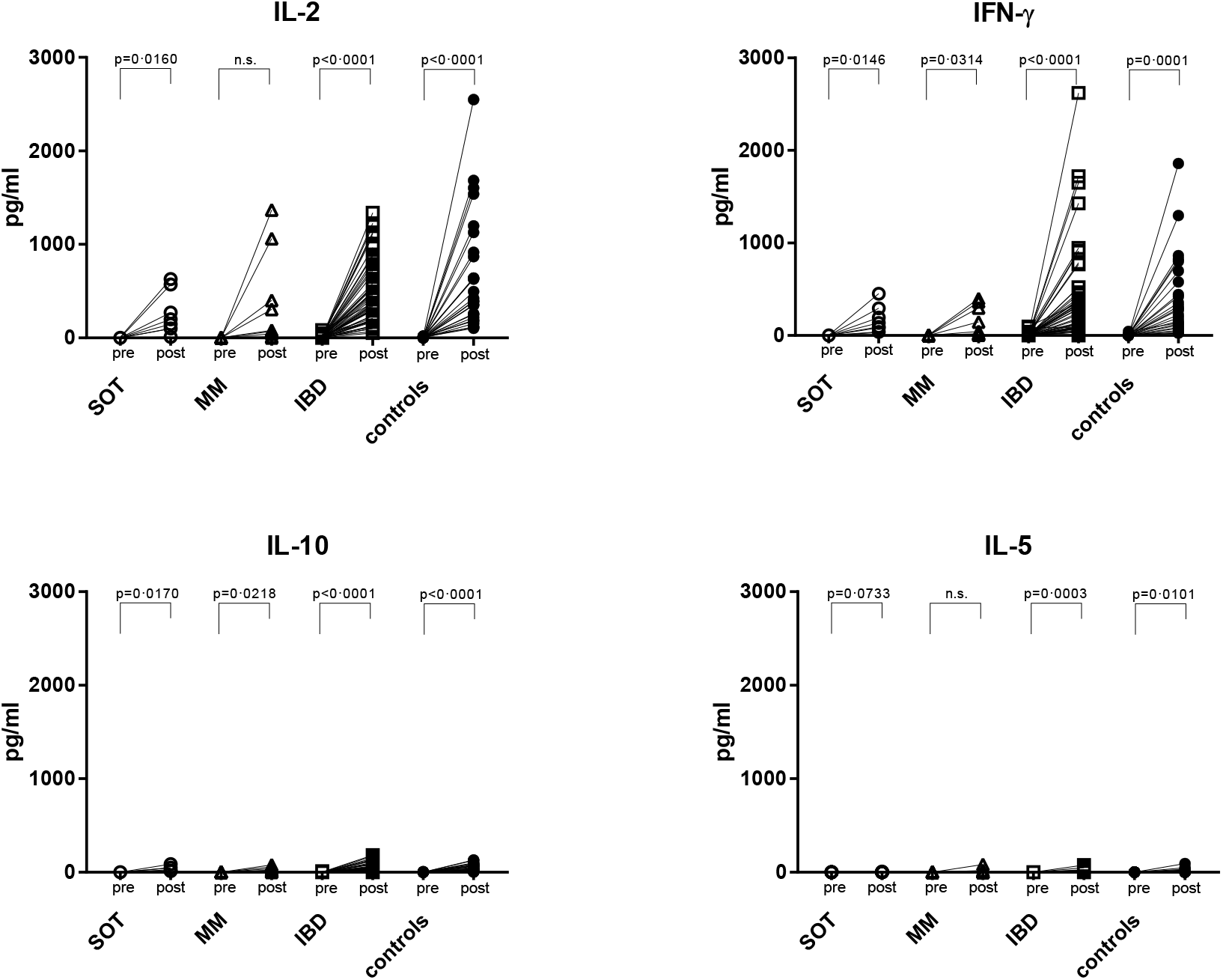
S1-spike induced cytokine production before vaccination and one week after the second mRNA vaccine dose. PBMCs were stimulated with the peptide pool of the S1 subunit of the SARS-CoV-2 spike protein for 24 hours. Afterwards, the supernatants were collected and IL-2, IFN-γ, IL-10 and IL-5 were measured by the Luminex system. SOT are represented as open circles, MM as open triangles, IBD as open squares and controls as full circles. Differences between the groups below p values of 0.05 were regarded as significant.

Further, in the controls and in IBD patients both the IL-2 (r^2^=0.1384, p=0.0072 and r^2^=0.2258, p=0.0141, respectively) and IFN-γ (r^2^=0.09839, p=0.0287 and r^2^=0.2193, p=0.0158, respectively) levels correlated positively with the antibody levels. With SOT and MM patients this was also true for IL-2 (r^2^=0.5039, p=0.0486 and r^2^=0.6627, p=0.0076, respectively) but not the case for IFN-γ (Fig 3; r^2^=0.00040, p=0.9624 and r^2^=0.3482, p=0.0943, respectively). Upon restimulation with the peptide pools derived from the SARS-CoV-2 nucleocapsid antigen, all cytokine levels remained at baseline levels after the second dose confirming that the immune response was derived from the mRNA vaccine, but not from a natural infection (Suppl Fig 5).

**Figure 3.**
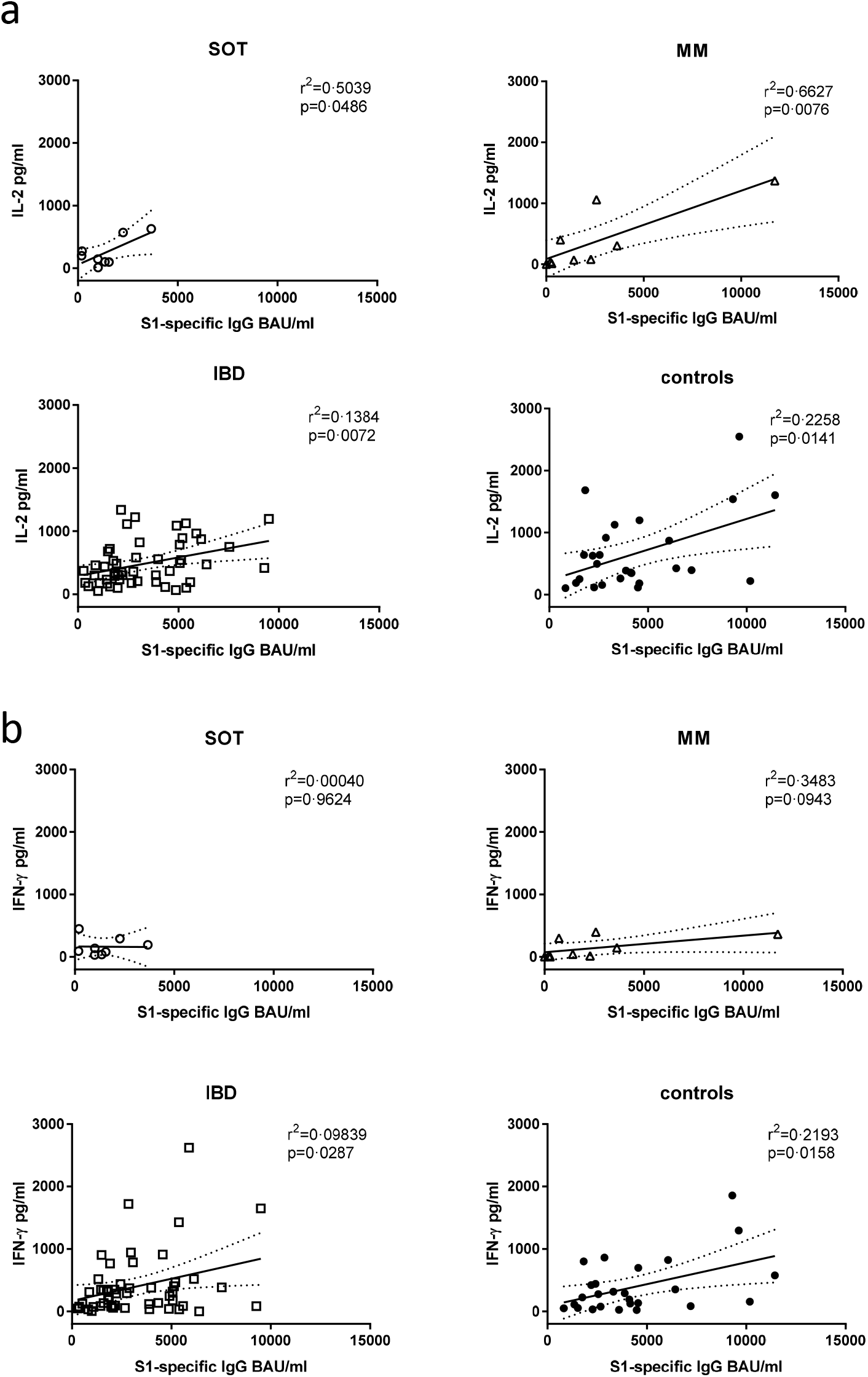
Correlation of antibody levels with cytokine production. The IL-2 (a) and IFN-γ (b) levels shown in Figure 2 were correlated with the S1-specific IgG antibody levels of week four after the second mRNA dose. Differences between the groups below p values of 0.05 were regarded as significant. Dotted lines represent 95% confidence intervals.

### Leukocyte phenotypes

Phenotyping of leukocytes was performed in whole blood samples in order to get an overview about the effects of the underlying diseases and the respective immunosuppressive drugs on cellular compartments.

Patients with MM displayed lower total absolute cell numbers of leukocytes, total lymphocytes, CD3^+^ T cells and CD3^+^CD4^+^ T-helper cells compared to controls (p=0.0398, p=0.0601, p=0.0342 and p=0.0017, respectively). With regard to CD19^+^ B cells, patients with SOT (p=0.0381) and MM (p=0.0073) had lower levels than controls. No significant differences were detected for monocytes, granulocyte, CD8^+^ T cells and NK cells, except for lower granulocyte counts in the MM patients compared to controls (p=0.0109) (Suppl Fig 6). In contrast to the cancer patients, we did not measure lower lymphocyte counts with IBD patients.

Correlation analysis showed that the B cell counts correlated with antibody levels at four weeks after the second dose in the total study population (Suppl Fig 7; r^2^=0.03777, p=0.0014).

### Persistence of humoral immune responses

Evaluation of the antibody levels five to six months after the second dose demonstrated that the peak levels measured after one month after the second dose had experienced a significant decline in all groups (p<0.0001) (Fig 5a), with SOT, MM and IBD patients exhibiting lower antibody titers (p=0.0108, p<0.0001 and p<0.0001, respectively) than the controls (Fig 5b). The most impressive mean fold-decrease was found in MM patients (12.9; p=0.0007) and in IBD patients (12.8; p<0.0001) followed by SOT patients (8.3; p=0.2413) compared to controls (6.0). With regard to IBD treatment, the decline rate was higher (p<0.001) in those with TNF-α inhibitors (46%) than without TNF-α inhibitors (33%). The highest percentage of study participants who turned seronegative five to six months after the second dose belonged to the MM group (19.6%) followed by 7.3% in SOT and 3.7% in IBD patients with any kind of treatment, whereas all controls remained seropositive (Fig 5c).

**Figure 4.**
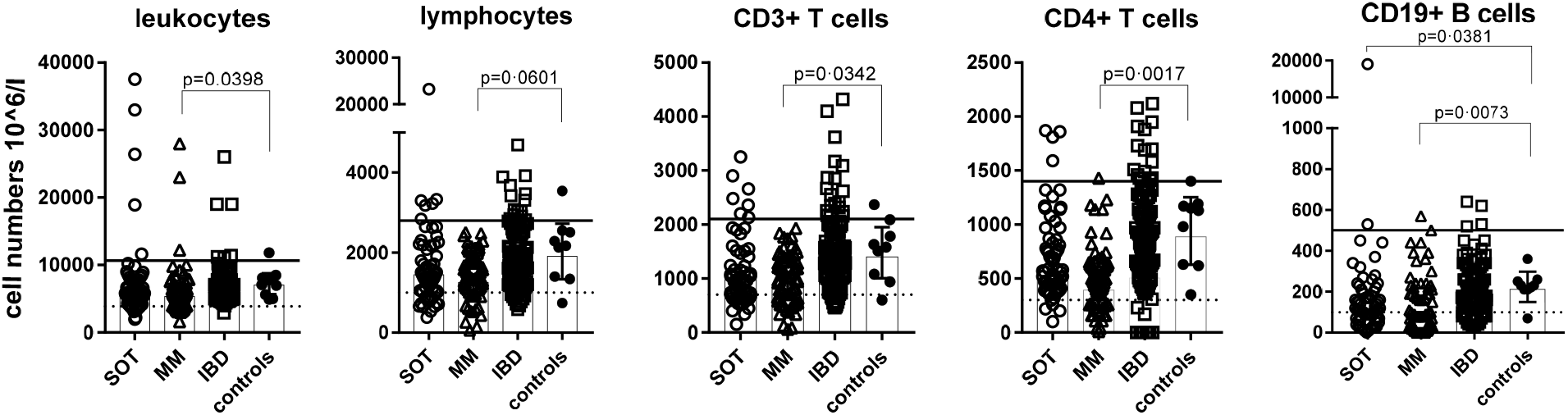
Typing of leukocytes. Total leukocytes, total lymphocytes, CD3^+^ and CD3^+^CD4^+^ T cells as well as CD19^+^ B cells are shown in absolute numbers (10^6/l) for all patients and nine controls; SOT are represented as open circles, MM as open triangles, IBD patients as open squares and controls as full circles. Leukocyte subtyping analysis was performed on the day of the first mRNA vaccine dose. Differences between the groups below p values of 0.05 were regarded as significant. Black lines indicate upper reference values and dotted lines indicate lower reference values. Columns represent GMC with 95% confidence interval (CI).

**Figure 5.**
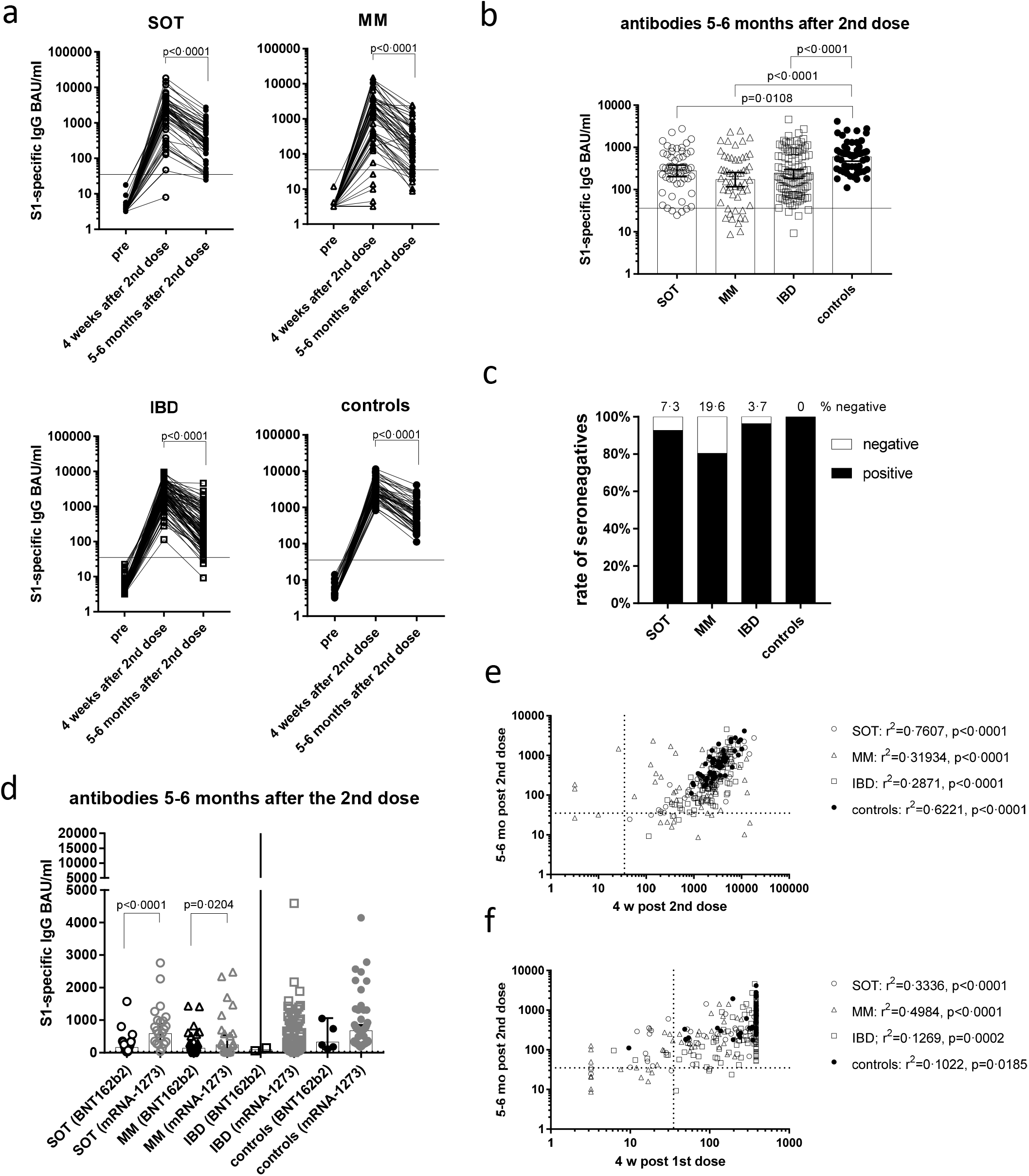
Persistence of antibody responses over a six months period. Antibody levels (a) were measured before the first vaccination and four weeks as well as five to six months after the second dose for all study groups. Individual S1-specific IgG levels (b) and % of seronegative participants (c) at five to six months after the second dose. S1-specific antibody levels five to six months after the second dose in respect of the mRNA vaccine applied (d). Correlation of the S1-specific IgG levels (in BAU/ml) four weeks and five to six months after the second mRNA (e), and correlation of the antibody levels four weeks after the first mRNA vaccine dose before receiving the second dose with those collected five to six months after the second dose (f). SOT are represented as open circles, MM as open triangles, IBD patients as open squares and controls as full circles. Differences between the groups below p values of 0.05 were regarded as significant. Dotted and black lines represent the threshold for positive results (35.2 BAU/ml). Columns represent GMC with 95% confidence interval (CI).

Cancer patients that received mRNA-1273 vaccination presented with higher seroconversion rates (Suppl Fig 8) and higher GMCs at months five to six, as compared to those vaccinated with BNT162b2 (581.2 vs. 163.5, p<0.0001 in SOT patients; 242.5 vs. 142.2, p=0.0204 in MM patients) (Fig 5d).

Furthermore, we found that the antibody levels measured four weeks after the second dose correlated positively with the level seen at 5 to 6 months in all groups (SOT: r^2^=0.7599, p<0.0001; MM: r^2^=0.3144, p<0.0001; IBD: r^2^=0.2548, p<0.0001; controls: r^2^=0.6189, p<0.0001) (Fig 5e). Even four weeks after the first dose and before the second dose, this positive correlation existed in cancer and IBD patients (SOT: r^2^=0.3325, p<0.0001; MM: r^2^=0.4939, p<0.0001; IBD: r^2^=0.14, p=0.0002; controls: r^2^=0.09411, p=0.0454). Thus, in the sera of MM patients who were seronegative after the first dose - but responded after the second - the antibodies quickly dropped below the threshold five to six months after the second dose (Fig 5f).

## Discussion

In the present study, we aimed to explore humoral and cellular immune responses after COVID-19 mRNA vaccination as well as the duration of antibody responses in immunocompromised patients with different disease entities and therapies compared to a healthy control group.

We show that vaccine-specific humoral and cellular responses to COVID-19 mRNA vaccines depend on disease entity and on the immunosuppressive drugs prescribed. The rate of non-responders according to a negative S1-specific ELISA after the first dose were highest in patients with MM (50%) and SOT (28.6%), but improved four weeks after the second dose to 17.1% and 1.6%, respectively. These results underline the importance of completing the two-dose schedule with mRNA vaccines and are in line with previous data showing diminished seroresponses within one month after the second dose in MM patients compared to controls.^14,15^ In addition, also patients with IBD and the controls showed a clear benefit from the second dose, as both groups contained a low rate of non-responders after the first vaccine dose (3.8% and 1.5%, respectively), but reached 100% seroconversion after the second dose. Whether the increase in seroconversion rate after the second dose was the effect of delayed antibody kinetics in cancer patients, as described for the elderly^16^, or a true effect of the second dose remains to be determined.

With regard to antibody levels, MM and SOT patients achieved lower antibody levels than IBD patients and the controls with the lowest levels in MM patients. Likewise, the most pronounced changes in lymphocyte counts were evident in MM patients with lymphopenia demonstrated by diminished overall CD3^+^ T cell numbers and particularly CD4^+^ T helper cells as well as CD19^+^ B cells. These changes are typical for multiple myeloma patients and their treatment^17^ and also explain the decreased seroresponses, since these cellular subsets are required for an effective immune response to be induced.^18^ These data suggest lymphocyte subtyping in these patient groups can be helpful to predict vaccine non-responsiveness before primary vaccination.

In both cancer patient groups (MM and SOT) stronger antibody responses were noted after the mRNA-1273 than after BNT162b2, most likely due to the higher mRNA content in mRNA-1273 (100 µg of the full dose) as compared to BNT162b2 (30 µg). Whether this observation may also indicate superior efficacy in patients with cancer is currently difficult to predict as a correlate of protection for SARS CoV-2 antibody levels is still not defined. Certain thresholds of antibody and neutralization titers have been published recently,^19,20^ but it still needs to be determined whether this threshold needs to be adapted with upcoming virus variants and the comparability of results expressed in BAU/ml and NT50 between different manufacturers and laboratories. Despite clear cut-off values for protection are still lacking and need to be harmonized, the determination of the quantity and quality of antibodies (WHO-benchmarked BAU values and neutralization titers) is already very useful to identify vaccine non-responders after the completion of a vaccination cycle (as was done in this study), and to immediately introduce these non-responders to booster vaccination, thus closing the phase of non-protection for these particularly vulnerable groups of people.

With regard to vaccine comparison in IBD patients and controls, only few were vaccinated with BNT162b2-compared to mRNA-1273 and therefore no conclusions or recommendations can be made for one of the mRNA vaccines.

When analyzing the magnitude of antibody levels in IBD patients in relation to their treatment regimens, only patients receiving TNF-α blockers exhibited lower antibody levels, but not those on other treatments such as the integrin antagonist vedolizumab or the IL-12/23 ustetkinumab. In this respect, there was no difference in antibody levels in patients receiving various drugs targeting TNF-α inhibition, which adds to a previous publication showing reduced GMT’s in infliximab treated patients compared to vedolizumab.^21^ JAK inhibitors are regarded as potent immunosuppressive drugs and the few IBD patients in our study on upatacitinib may indicate a low seroconversion rate. Along these lines, a recent study demonstrated non-responder rates of up to 33% in arthritis patients on the JAK inhibitor upatacitinib.^22^

Vaccine-specific T cell reactivity to S1 measured by cytokine secretion was induced in all groups after two vaccine doses. However, in the MM and SOT groups the levels were low in comparison to the IBD patients and the controls. In general, the levels of IFN-γ and IL-2 correlated well with the S1-specific antibody levels, meaning that high antibody levels are associated with positive cytokine levels and vice versa. However, a high cellular response seems not to be a prerequisite for the formation of antibodies, because some individuals revealing only marginal cytokine induction did mount an antibody response well above the threshold for positivity after the second vaccine dose. This dissociation between cellular and humoral responses was noticed particularly in SOT patients of our study. Differences in cellular and humoral responses were also reported in a recent publication on vaccine responses after two doses of BNT162b2 in nursing home residents^23^ as well as in a study with rheumatoid arthritis patients treated with anti-CD20 antibodies that did not mount antibody production but significant IFN-γ levels.^24^

Regarding cellular responses in antibody-non-responders, we had only one non-responder in our preselected subgroup, whose T cell responses were analyzed. This individual did not mount any cytokine response either, a phenomenon also known in response to other vaccines.^25^ However, in antibody-non-responders there remains the possibility that in cases of cytokine responses even at low levels the generation of memory T cells might contribute to development of protective immune responses upon contact with the natural virus. In this respect, the production of IFN-γ seems to be of importance, since this has been linked with a less severe course of COVID-19 during early infection.^26^ Along these lines, it can be of interest to analyze T cell responses in antibody-non-responders to guide the decision regarding additional (i.e. third or fourth) vaccine doses in immunocompromised individuals, although it must be kept in mind that an interpretation of T cell responses and their correlation with protection from COVID-19 is awaiting confirmation.

Concerning the persistence of antibody levels, we found antibody peak levels around four weeks after the second mRNA dose and after five to six months in all groups. Though, in several MM patients the antibody levels fell under the threshold prior to the six months interval after the second dose emphasizing that in immunocompromised risk groups a third dose needs to be offered earlier. A study in organ transplant recipients already showed that a third COVID-19 vaccine dose two months after the second vaccine dose increased the rate of seroconversion and further increased already existing antibody titers.^27^ In this context, a third dose in severely immunosuppressed individuals already four weeks after the second dose has now been licensed for both vaccines, BNT162b2 and mRNA-1273. Of note, antibody levels were maintained at higher levels after five to six months in SOT and MM patients, rather upon administration of mRNA-1273 than of BNT162b2.

All control participants and even the elderly maintained positive antibody titers that can be considered protective along with high cytokine levels for at least six months as compared to the other investigated groups, indicating that booster vaccination would not be needed as early as six months after the second vaccination. However, caution may be needed for elderly populations, as it was shown recently that up to 40% of the elderly had lost detectable antibody levels at six months^28^ which also helps to explain increasing breakthrough infections in this population^29^ and thus supporting the earlier administration of a third vaccine dose particularly in healthy elderly. With regard to emerging SARS-CoV-2 variants of concern, a clearly reduced neutralizing activity of vaccine-induced antibodies against the delta variant has been observed,^30^ which might argue for rather timely booster vaccinations also in healthy vaccinees.

In conclusion, our data clearly shows that a third dose of mRNA vaccine is needed for immunocompromised patients already before six months after the second dose. Hence, earlier vaccination should be discussed more broadly even for non-severely immunosuppressed individuals, who lack adequate antibody production. Therefore, we suggest antibody testing for immunocompromised individuals to identify non-responders and help to communicate the additional use of non-pharmaceutical measures. Additionally, our data suggest that the vaccination with mRNA-1273 could be more immunogenic in immunocompromised patients. For antibody non-responders, the application of further vaccine doses (or the prophylactic application of monoclonal anti-SARS-CoV-2 antibodies) may be an option for some individuals, particularly when vaccine-specific cellular responses are elicited.

## Data Availability

Upon reasonable request and depending on a positive ethics vote data can be provided by the principal investigator.

## Authors contributions

Literature search: AWa, ST, CZ, GN, HS, WR, UW; figures: AWa, ET; study design: AWa, ST, WR, UW; data collection: AWa, EGS, AS, MO, BK, WFP, AOR, GT, LG, AWe, IZ ET, JJ, UW; data analysis: AWa, AS, AOR, GT, LG, BK, WFP, EGS, ET, MK, HS, UW; data interpretation: AW, EGS, AOR, GT, LG, ET, HS, GN, WR, CZ, UW; writing: AW, CZ, UW; Revising the manuscript: AW, EGS, AS, MO, ET, AW, IZ, AR, LG, BK, MK, ST, WFP, MK, HS, GN, WR, CZ, UW.

## Acknowledgements

We would like to thank the clinical study team Dooa Al-Mamoori, Lisa Dohr-Loufouma, Peter Pichler, Melita Poturica, Andrea Schagerl and Claudia Seidl-Friedrich for their efforts at the Institute of Specific Prophylaxis and Topical Medicine. Furthermore, we would like to thank Petra Waidhofer-Söllner for her expertise in measuring cytokines, Ulrike Körmözci, and Theresa Oberhofer and Arno Rottal for their help with lymphocyte subtyping at the Institute of Immunology. We are grateful for the commitment of the serology team Tatjana Matschi, Vanessa Maurer, Barbara Schaar, Karin Schoiswohl, Andrea Wendl to perform all antibody measurements at the Institute for Hygiene and Applied Immunology of the Medical University of Vienna.

We thank all the patients for participating in the trial and would like to thank the myeloma groups, particularly Mrs. Jirsa and Mrs. Pearsall.

## Data availability

Ethics approval does not allow data sharing of person related data. Upon reasonable request and depending on a positive ethics vote data can be provided by the principal investigator.

