## Supplementary Material and Figures for "Humoral and cellular immune responses and their kinetics vary in dependence of diagnosis and treatment in immunocompromised patients upon COVID-19 mRNA vaccination"

**Table of contents**

Supplementary materials page 2

Supplemantary Figures

Supplementary Figure 1 page 4
Supplementary Figure 2 page 5
Supplementary Figure 3 page 6
Supplementary Figure 4 page 7
Supplementary Figure 5 page 8
Supplementary Figure 6 page 9
Supplementary Figure 7 page 10
Supplementary Figure 8 page 11

Study population

We invited patients without prior COVID-19 vaccination suffering from solid tumors (SOT), multiple myeloma (MM), inflammatory bowel disease (IBD) as well as healthy individuals (controls) via the outpatient vaccination clinic at the Institute of Specific Prophylaxis and Tropical Medicine and the Department of Internal Medicine of the Medical University Vienna and General Hospital of Vienna to participate in this study. In total, 361 participants were enrolled and 329 with various immunosuppressive/-modulatory treatments were included for the final analysis, excluding those participants with prior COVID-19 infection, with incomplete two-dose mRNA vaccination schedule or lost to follow-up (Suppl Fig 1 and 2). All 329 participants received two doses of a mRNA vaccine (either BNT1622b2, Comirnaty, Pfizer BioNTech or mRNA-1273, Spikevax, Moderna Biotech) administered in intervals of three and four weeks, respectively. Serum samples for antibody titers measurements were collected before the first dose, on the day of the second dose and four weeks as well as five to six months after the second dose. Sera were frozen and stored until analysis. Participants without antibody responses four weeks after the second dose (SOT: n=1; MM: n=12) were offered an earlier third dose and therefore were not further included in the analysis of antibody responses five to six months after the second dose. In a subgroup (n=102) and upon participants consent lithium heparinized blood was taken before the first dose and seven days after the second dose to isolate peripheral blood mononuclear cells (PBMC).

T-cell responses

For each participant, we seeded 190 µl medium containing 5 x $10^5$ of the freshly isolated PBMCs into each of four wells of a 96-well plate (Cellstar Cat.-No.: 650 180, Greiner Bio-One GmbH, Kremsmünster, Austria). The cultivation medium used was RPMI 1640 supplemented with 2 mM L-glutamine (Lonza™, BioWhittaker™, Fisher Scientific, Vienna, Austria) and 5% human AB serum (PAN-Biotech Seraclot, Aidenbach, Germany). The cells were rested over night before adding the stimulants. Two different SARS-CoV-2 specific stimulants were used, namely the PepTivator® SARS-CoV-2 peptide pools of the S1 domain of the spike protein and of the nucleocapsid protein N (Cat. No. 130-127-041 and 130-126-698) Miltenyi Biotech, Bergisch Gladbach, Germany). The lyophilized antigens were reconstituted in sterile water (Cat. No. 95284, Merck, Darmstadt, Germany) as described in the manufacturer's instructions yielding a stock concentration of 30 nmol of each peptide per ml. Working aliquots with a concentration of 3 nmol of each peptide per ml in 1 x DPBS (Gibco™ Cat. No. 14190094, Thermo Fisher Scientific, Vienna, Austria) were prepared and stored at -80°C until usage. Rested PBMC were stimulated with 10 µl of these working solutions resulting in a stimulation with 0·03 nmol per peptide per 5 x $10^5$ cells in a final volume of 200 µl. In parallel, stimulation with 10 µl of 1 x DPBS or with 10 µl of PHA (100 µg/ml working solution; 5 µg/ml final concentration) was performed as a negative or positive control, respectively. The cells were stimulated for 24 hours after which the supernatant was taken and stored at -80°C until cytokine measurement. For the latter, we used the Luminex® 100/200 System and determined the concentration of the following cytokines: interleukin (IL)-2, IL-5, IL-10, IL-17a, IL-22, granulocyte-macrophage colony-stimulating factor (GM-CSF) and interferon (IFN)-γ using Luminex Human High-Sensitive Cytokine Performance Assays from Bio-Techne (Minneapolis, MN, USA) according to the manufacturer's instructions.

Neutralizing antibodies

Neutralization assay was performed in the preselected subgroup of patients where cellular responses were evaluated and in all ELISA-negative participants with the human SARS-CoV-2 isolate BetaCoV/Munich/BavPat1/2020, which was kindly provided by Christian Drosten, Charité, Berlin, and distributed by the European Virology Archive (Ref-SKU: 026V-03883).[1] The virus was passaged once through the human lung adenocarcinoma cell line Calu-3 (ATCC HTB-55™, kindly provided by Walter Berger, Medical University of Vienna, Vienna, Austria) to obtain a high titer virus stock. Neutralization assay was performed according to the protocol by Amanat et al..[2] Briefly, one day prior the assay, 10000 Vero cells (ATCC CCL-81™, kindly provided by Sylvia Knapp, Medical University of Vienna, Vienna, Austria) were seeded into each well of a 96-well plate (in a Dulbecco´s Modified Eagle´s medium, DMEM, Gibco/Thermo Fisher, high glucose, with GlutaMAX and sodium pyruvate, supplemented with 10% fetal calf serum, FCS, Biowest, Nuaillé, France; 1% MEM non-essential amino acids solution, Gibco/Thermo Fisher; 100 U/ml penicillin and 100 µg/ml streptomycin, Gibco/Thermo Fisher). On the next day, paired pre-immune and immune heat-inactivated (56°C for 30 min) proband sera were serially diluted in DMEM medium with reduced serum (2% FCS) and incubated in duplicates with the SARS-CoV-2 virus (80 µl, equaling 800 half-maximal tissue culture infectious dose (TCID50) per well; final volume 160 µl per well) for 1 hour at 37 °C in a Biosafety Level 3 facility of the Medical University of Vienna. After one hour, 120 µl of the mixture was used to infect the monolayers of Vero cells, achieving infection of 600 TCID50/well.

48 hours later, infected cells were fixed with 10% formaldehyde in PBS, followed by 5% formaldehyde postfix. Subsequently, cells were washed with PBS, permeabilized with 0·1% Triton X-100 detergent in PBS, blocked with a blocking buffer (10% FCS in PBS+0·05% Tween-20 detergent), and stained with a rabbit anti-SARS-CoV-2 nucleocapsid monoclonal antibody (40143-R019, SinoBiological, Beijing, China) diluted 1:15000 in blocking buffer), followed by the goat anti-rabbit-HRP antibody (170-6515, Bio-Rad, diluted 1:10000 in blocking buffer). In-Cell ELISA was then developed using DY999 substrate solution according to the manufacturer's recommendations (R&D Systems, Minneapolis, MN, USA) and measured at 450 nm (and at 630 nm to assess the background) using a Mithras multimode plate reader (Berthold Technologies, Bad Wildbad, Germany). To calculate percent neutralization at each well, the following formula was used: 100 − [(X - average of 'no virus' wells)/(average of 'virus only' wells - average of 'no virus' wells)*100], where X is the background-subtracted read for each well. Titers were calculated in GraphPad Prism 9 by generating a 4-parameter logistical fit of the percent neutralization at each serial serum dilution. The 50% virus neutralization titer (NT50) was reported as the interpolated reciprocal of the dilution yielding a 50% reduction in the anti-SARS-CoV-2 nucleocapsid staining.

**Suppl Figure 1. Flow chart.** Description of participants included in the study.

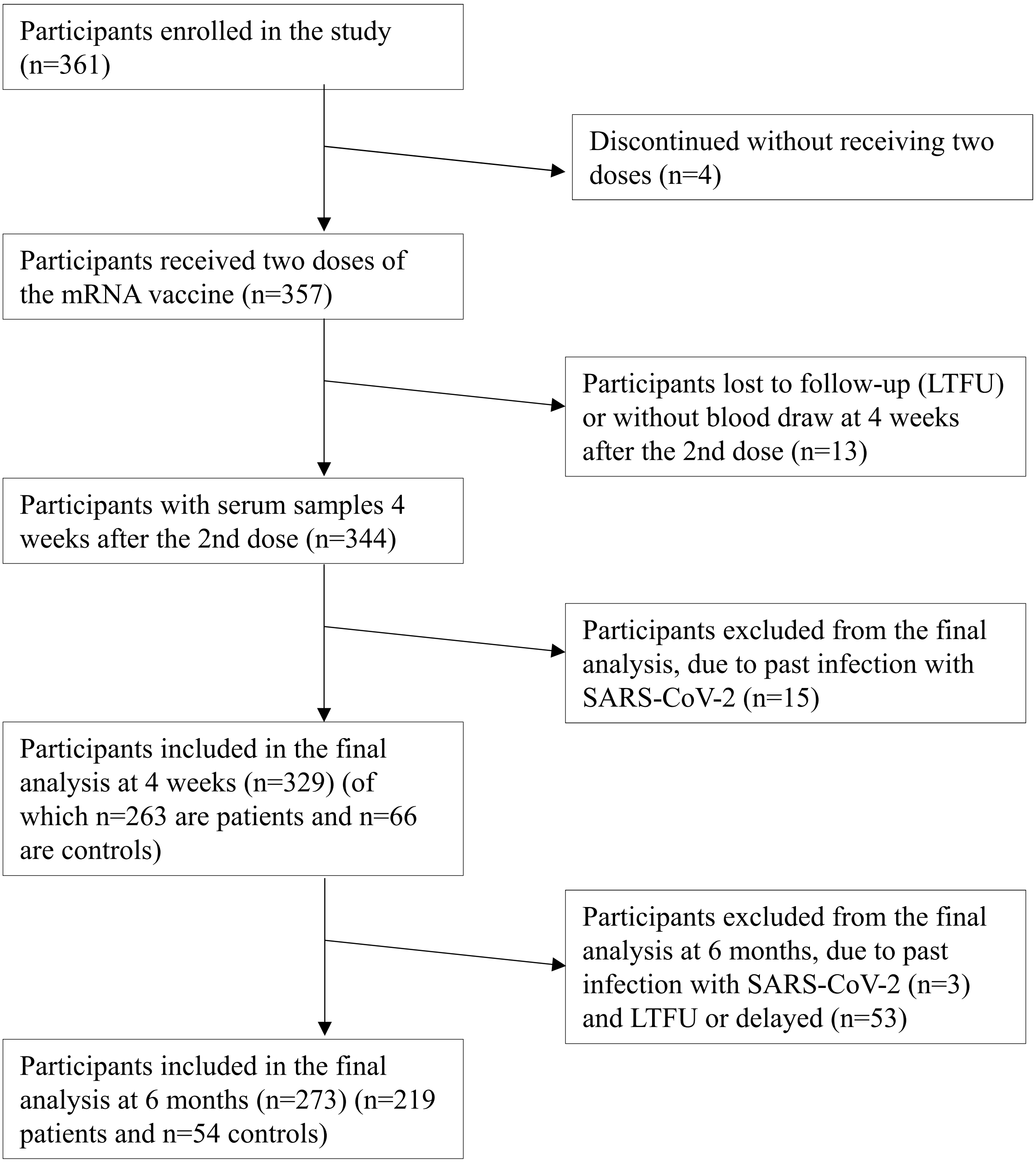

**Suppl Figure 2. Ongoing treatment in patients (SOT, MM and IBD).** Percentages of different treatment compounds prescribed in the cancer and IBD patients with ongoing treatment.

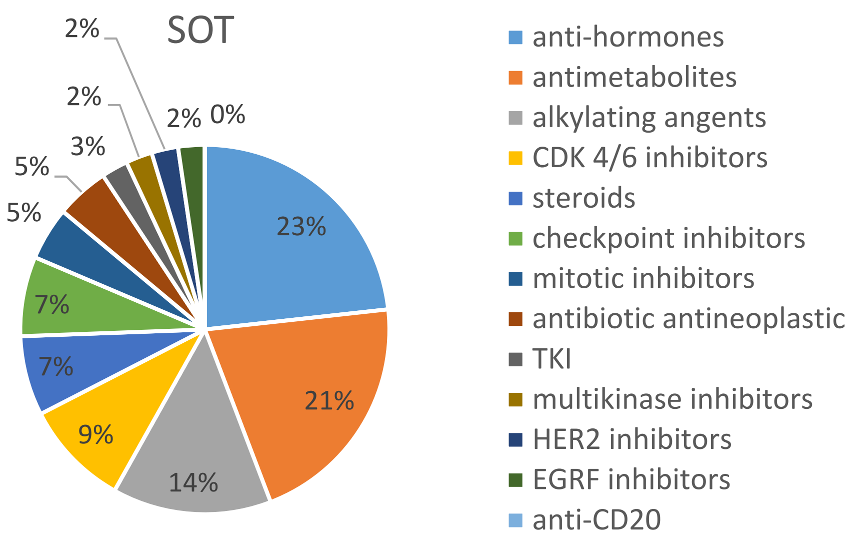


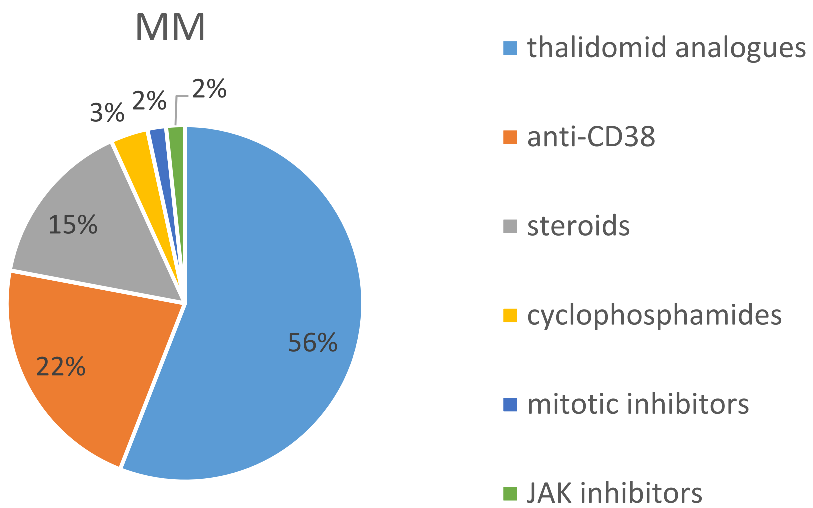


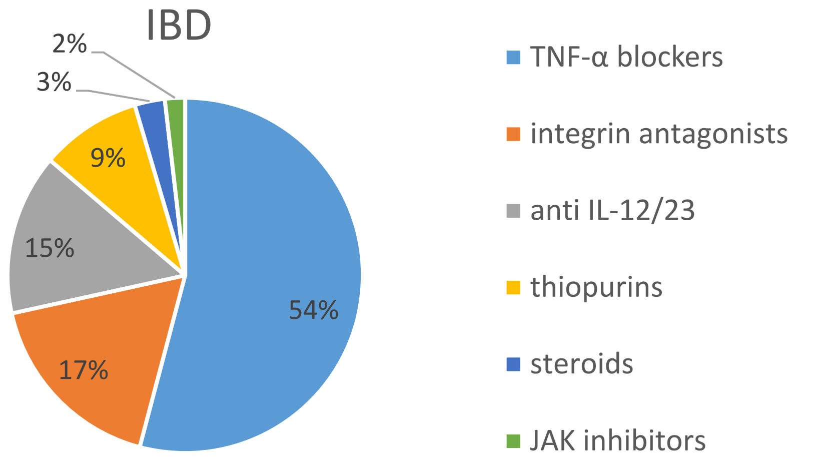

**Suppl Figure 3. Antibody responses four weeks after the second mRNA vaccination in relation to age, gender and neutralization capacity.** Individual S1-specific IgG antibodies in relation to age (a) and gender (b). Neutralization titers before the first and after the second mRNA vaccine dose (c). Correlation of BAU/ml and NT50 with ELISA-negative patients (n=13) (d). SOT are represented as circles, MM as triangles, IBD patients as squares and controls as full black or grey circles. Dotted lines indicate the threshold for positive results (35·2 BAU/ml and NT50 1:76) and black line represents the limit of detection (<3·2 BAU/ml) in the ELISA. Differences between the groups below p values of 0·05 were regarded as significant. Columns represent GMC with 95% confidence interval (CI).

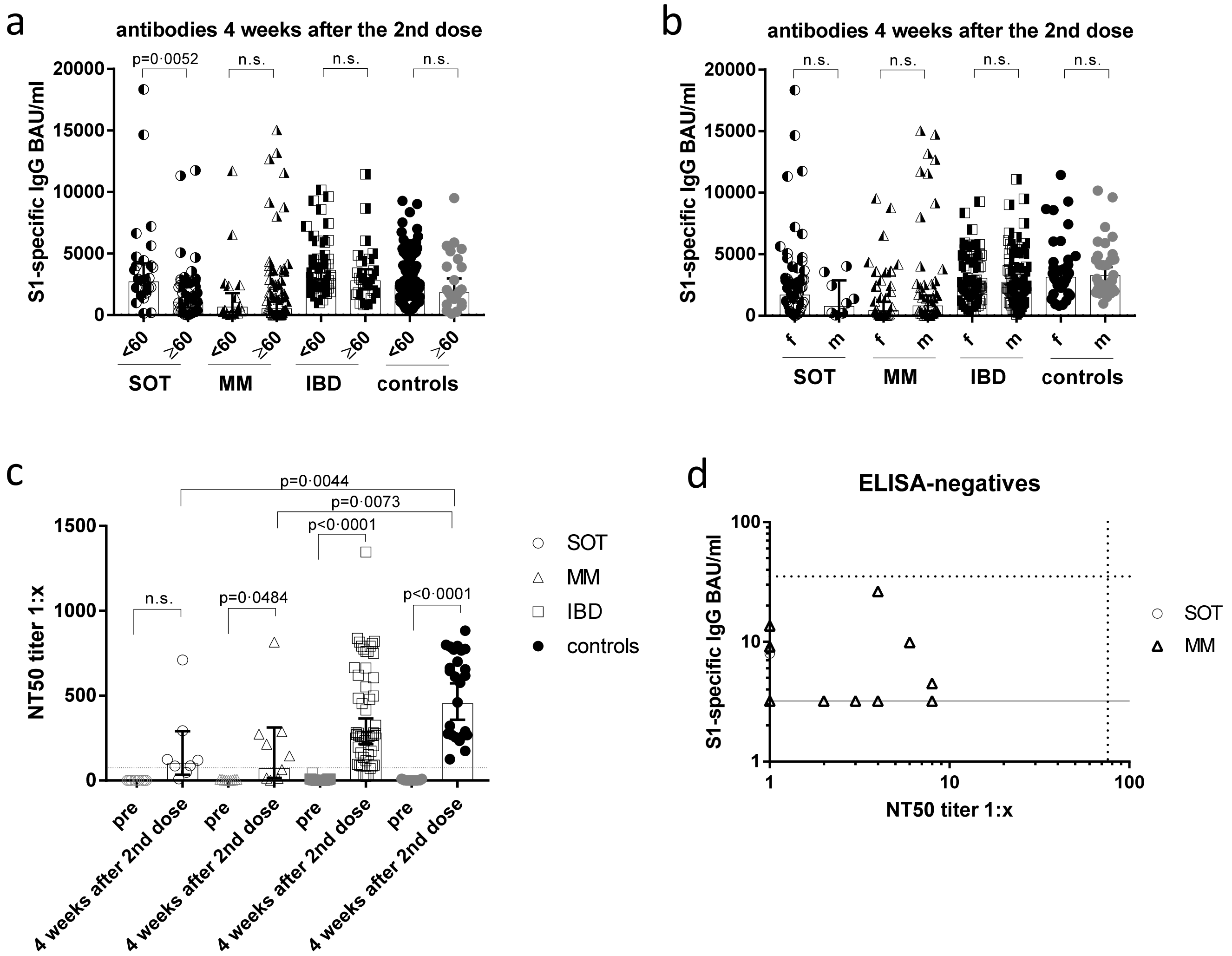

**Suppl Figure 4. S1-spike induced cytokine production before vaccination and one week after the second dose of the mRNA vaccine.** PBMCs were stimulation with the peptide pool of the S1 subunit of the SARS-CoV-2 spike protein for 24 hours. Afterwards, the supernatants were collected and GM-CSF, IL-17a and IL-22 were measured by the Luminex system. SOT are represented as open circles, MM as open triangles, IBD as open squares and controls as full circles. Differences between the groups below p values of 0·05 were regarded as significant.

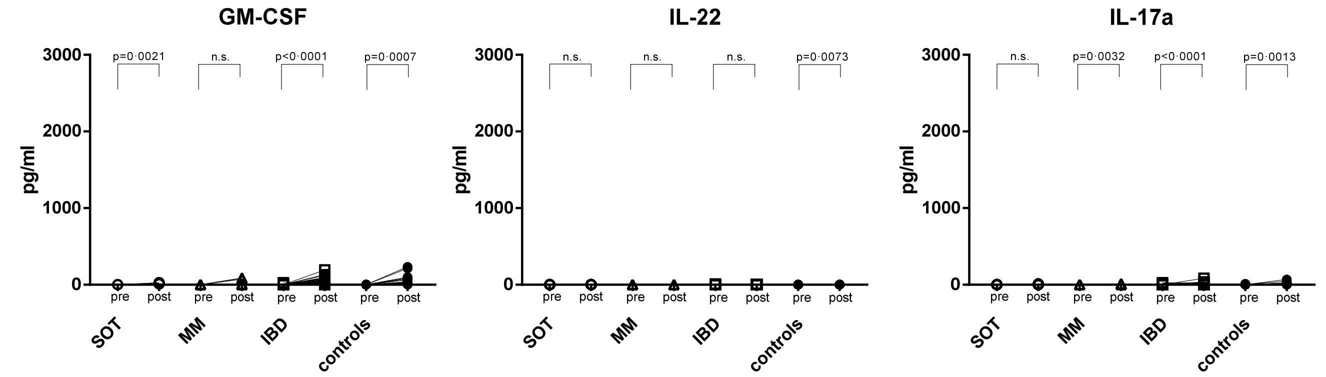

**Suppl Figure 5. Nucleocapsid induced cytokine production before vaccination and one week after the second dose of the mRNA vaccine.** PBMCs were stimulation with the peptide pools of the nucleocapsid protein of SARS-CoV-2 for 24 hours. Afterwards, the supernatants were collected and IL-2, IFN-g, IL-10, IL-5, GM-CSF, IL-17a and IL-22 were measured by the Luminex system. SOT are represented as open circles, MM as open triangles, IBD as open squares and controls as full circles. Differences between the groups below p values of 0·05 were regarded as significant.

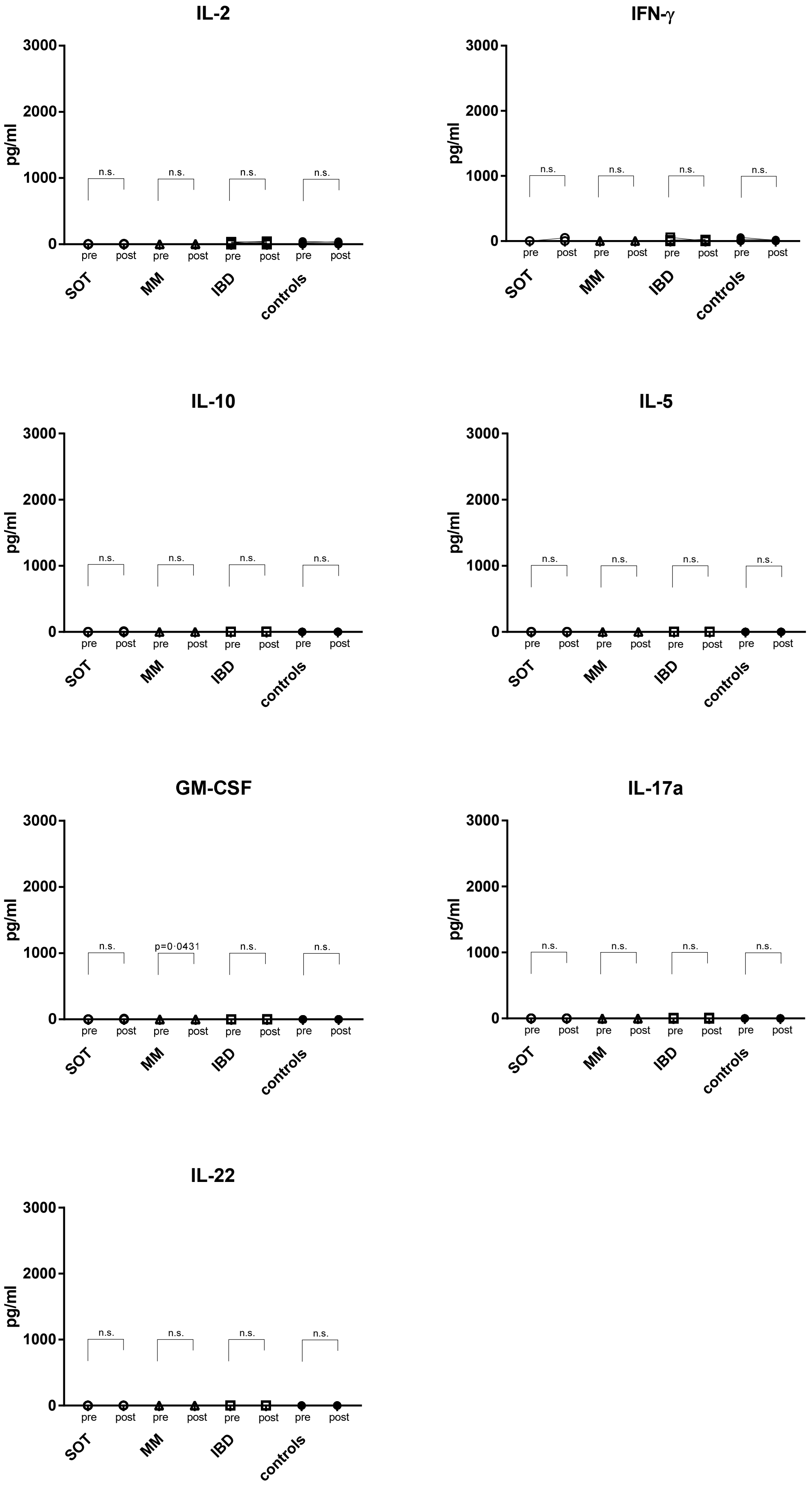

**Suppl Figure 6. Typing of leukocytes.** Monocytes, granulocytes, $CD8^{+}$ T cells and NK cells are shown in absolute numbers (10^6/l) for all patients and nine controls; SOT open circles, MM open triangles, IBD open squares and controls full circles. Leukocyte subtyping analysis was performed on the day of the first mRNA vaccine dose. Differences between the groups below p values of 0·05 were regarded as significant. Black lines indicate upper reference values and dotted lines indicate lower reference values. Columns represent GMC with 95% confidence interval (CI).

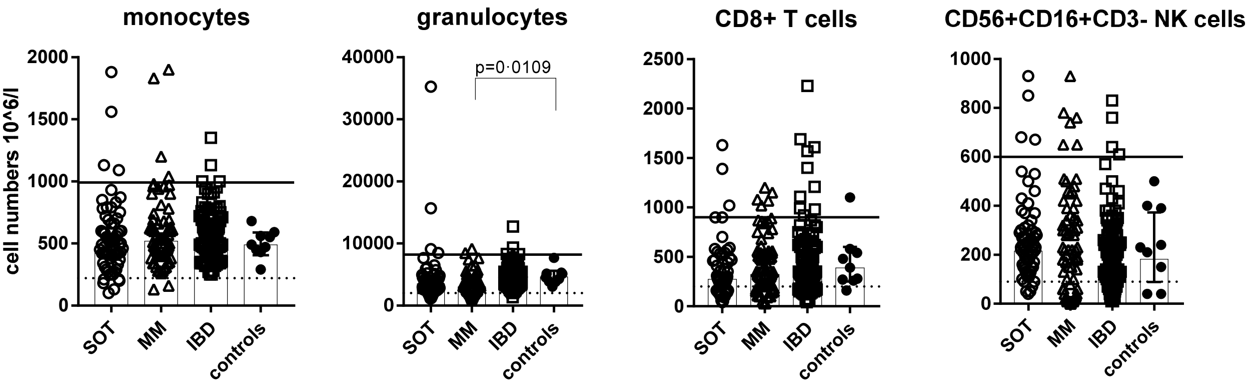

**Suppl Figure 7. Correlation of antibody production with CD19⁺ B cell counts.** S1-specific IgG levels measured 4 weeks after the second mRNA vaccine dose were correlated with the B cell counts measured on the day of the first vaccine dose. Dotted lines represent 95% confidence intervals.

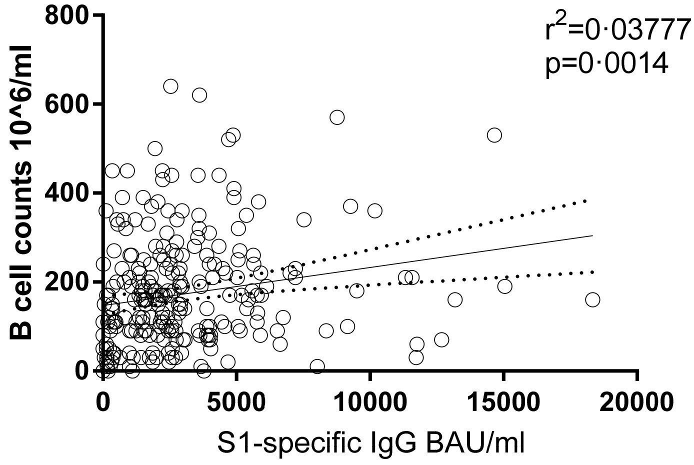

**Suppl Figure 8. Rate of seronegative participants in relation to the mRNA vaccine applied.** Seroconversion rates are shown four weeks after the first mRNA vaccination before receiving the second dose, and four weeks and five to six months after the second dose in SOT and MM participants after either BNT162b2 or mRNA-1273 vaccination.

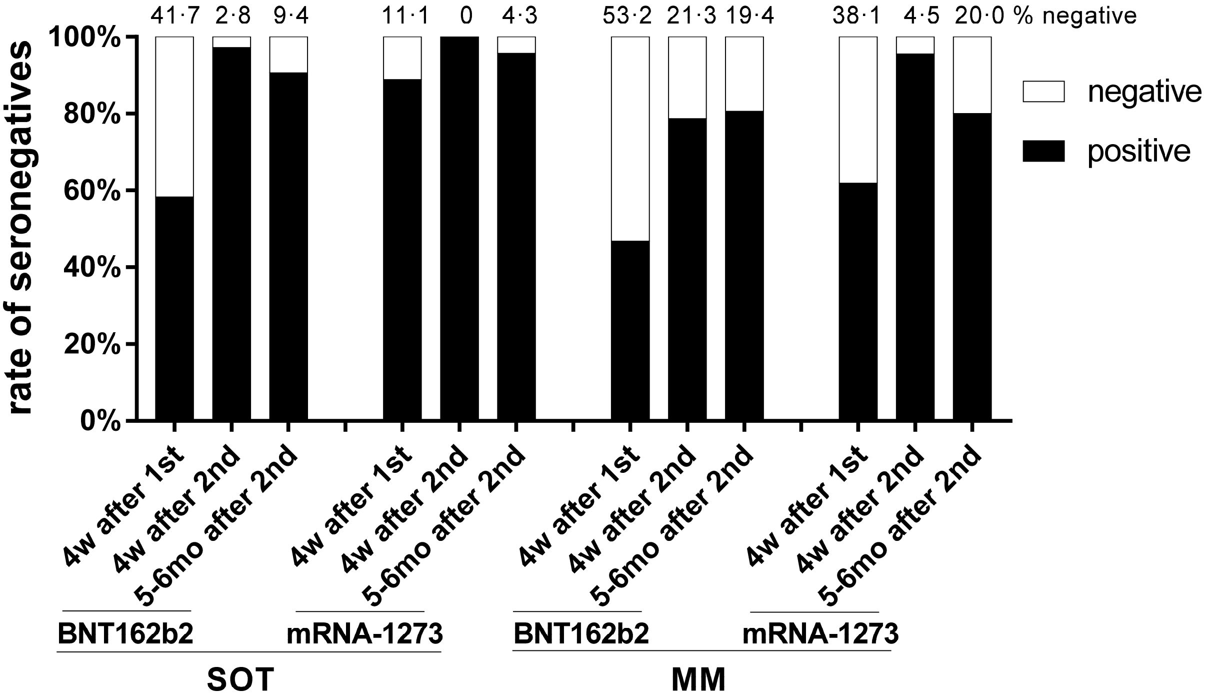
